# Estimating social contact rates for the COVID-19 pandemic using Google mobility and pre-pandemic contact surveys

**DOI:** 10.1101/2023.12.19.23300209

**Authors:** Em Prestige, Pietro Coletti, Jantien Backer, Nicholas G. Davies, W. John Edmunds, Christopher I. Jarvis

**Affiliations:** Centre for Mathematical Modelling of Infectious Diseases, London School of Hygiene & Tropical Medicine, United Kingdom; Data Science Institute, I-BioStat, Hasselt University, Hasselt, Belgium; National Institute for Public Health and the Environment (RIVM), Bilthoven, The Netherlands

**Keywords:** social mobility data, social contact rates, COVID-19 pandemic, prediction methods

## Abstract

During the COVID-19 pandemic, aggregated mobility data was frequently used to estimate changing social contact rates. By taking pre-pandemic contact matrices, and transforming these using pandemic-era mobility data, infectious disease modellers attempted to predict the effect of large-scale behavioural changes on contact rates. This study explores the most accurate method for this transformation, using pandemic-era contact surveys as ground truth. We compared four methods for scaling synthetic contact matrices: two using fitted regression models and two using “naïve” mobility or mobility squared models. The regression models were fitted using the CoMix contact survey and Google mobility data from the UK over March 2020 – March 2021. The four models were then used to scale synthetic contact matrices—a representation of pre-pandemic behaviour—using mobility data from the UK, Belgium and the Netherlands to predict the number of contacts expected in “work” and “other” settings for a given mobility level. We then compared partial reproduction numbers estimated from the four models with those calculated directly from CoMix contact matrices across the three countries. The accuracy of each model was assessed using root mean squared error. The fitted regression models had substantially more accurate predictions than the naïve models, even when models were applied to out-of-sample data from the UK, Belgium and the Netherlands. Across all countries investigated, the linear fitted regression model was the most accurate and the naïve model using mobility alone was the least accurate. When attempting to estimate social contact rates during a pandemic without the resources available to conduct contact surveys, using a model fitted to data from another pandemic context is likely to be an improvement over using a “naïve” model based on mobility data alone. If a naïve model is to be used, mobility squared may be a better predictor of contact rates than mobility per se.

## 1. Introduction

The COVID-19 pandemic caused extraordinary changes in social behaviour. Individuals reduced their social contact both spontaneously in response to perceived risk and following the imposition of physical distancing measures and restrictions on movement. The need to estimate the effect of these behavioural changes on viral transmission motivated the deployment of large-scale contact surveys and the collection of aggregated mobility data across the world.

Contact surveys measure the rates at which people come into physical or conversational contact with others, typically over a 24-hour period. One such survey was the CoMix survey (1) which began in the UK (2), Belgium (3) and the Netherlands (4) in 2020 and collected data from multiple countries in Europe throughout the pandemic (5). Contact surveys can be used to parameterise infectious disease models which take social contact rates as an input (6,7). However, they are expensive and difficult to undertake, so data was collected only in a limited number of countries. In early 2020, several companies started to release aggregated “mobility” data with the stated aim of helping public health professionals to understand behavioural change in response to the pandemic, as mobility is considered a valid proxy of risk behaviour (8). Usually, this mobility data quantified the time spent in different locations by mobile phone users, was made freely available, and covered participants in nearly all countries. These advantages led to the heavy use of mobility data during the pandemic to assess the impact of social distancing measures and to parameterise mathematical models, with Google’s “Community Mobility Reports” (9) being particularly widely used (10–13). However, as the use of mobility data as a proxy for social contact rates has not been formally assessed, it remains unclear how precisely to transform relative changes in mobility in different locations into changes in contact rates, and how well such transformations predict measured contact rates.

In this study, we explore methods for using aggregated mobility data to estimate social contact rates during a pandemic. Each method starts with a synthetic contact matrix that captures age-specific rates of social mixing in a pre-pandemic context, and scales the matrix using time-varying mobility data. We compare four methods for scaling synthetic contact matrices, two using fitted regression models with UK data and two using “naïve” mobility or mobility squared models, and determine which method best approximates empirical contact matrices collected in the CoMix study for out-of-sample data for the UK, Belgium, and the Netherlands. Our study helps inform modelling approaches where mobility data is used as a proxy for social contact behaviour and detailed social contact surveys are not available.

## 2. Materials and Methods

In this study, we visually compare trends in mean contacts recorded in the CoMix social contact survey with Google mobility data, and formally assess the performance of different methods used to relate mobility to social contacts. Specifically, we investigated the relationship between specific Google mobility indicators and corresponding contact types from the CoMix dataset for the UK, and developed a series of statistical and mechanistic models to relate the data to one another. We then used these models to estimate contact rates from mobility data for the UK, Belgium, and the Netherlands, in order to estimate the predictive accuracy of each model. Our results serve as a tool for epidemiologists and infectious disease modellers to understand better how mobility data might relate to contact rates in a population.

### 2.1 Study Design

The study design and method of informed consent for the CoMix study were approved by the Ethics Committee of the London School of Hygiene and Tropical Medicine (reference number 21795) in the UK and by the Ethics Committee of Antwerp University Hospital (reference 3236 - BUN B3002020000054) in Belgium. Need for approval was waived by the Medical Research Ethics Committee NedMec in the Netherlands (research protocol number 22/917). New approvals for use of all CoMix data, for the UK, Belgium and the Netherlands, was not necessary as secondary analyses were covered in the original approvals. All analyses were carried out on anonymised participant data. Additionally, Google mobility data was obtained from a publicly available source (9).

#### 2.1.1 Mobility data

Google’s Community Mobility Reports data uses the median value from a 5-week baseline period of 02/01/2020 to 06/02/2020 to compare changes in the number of visits to specific services/areas. The raw data is expressed as a percentage change relative to the baseline, e.g. -50% for half as many daily visits as during the baseline period and +100% for twice as many daily visits as during the baseline period. We transformed this to a scale which expresses the change from baseline as a multiple, e.g. 0.5 for half as many visits as compared to baseline and 2 for twice as many visits as compared to baseline. We included the Google mobility indicators for “retail and recreation”, “grocery and pharmacy”, “transit stations”, and “workplaces” in our analyses. Data is available between 23/03/2020 to 13/10/2022. We were unable to find clear information in regard to how Google classified visits into the preceding categories (14).

#### 2.1.2 Survey data

The CoMix survey collected information on contacts weekly in the UK from 23/03/2020 to 01/03/2022 (1). The present study includes weekly survey results for the entire study period for the visual comparisons between CoMix data and mobility indicators and for the descriptive statistics of survey participants; subsequent statistical models were created using CoMix and mobility data for the UK limited to between 23/03/2020 to 31/03/2021 as correlations between mean contact numbers and mobility became weaker in the second year (see supplementary Table S5.1). Adult panels at first contained 1500 participants, and increased to include 2500 participants from August 2020. Participants from each panel were surveyed once every 2 weeks, with panels alternating so that each week was covered. Ipsos MORI used quota sampling, for age, gender, and region, to recruit a representative sample of the UK (15). The survey followed the design of the 2005/2006 POLYMOD survey (16) with some additional questions. More details on the CoMix survey can be found in Gimma et al. and Jarvis et al. (2,15). CoMix data for out-of-sample tests of the regression model, UK (01/04/2021-01/03/2022); Belgium (16/04/2020-01/04/2021); the Netherlands (16/04/2020-01/04/2021), are detailed elsewhere (17–19). Survey participants were asked to report the number of people they met on the day prior to the survey in various settings. Participants had the option of recording any of their contacts in one of two ways: either individually (i.e. reporting details about contact made with one individual), or as mass contacts (i.e. reporting a summary of contact made with a group of several people). We processed these two contact types differently, as detailed below.

Traditionally, social contact surveys report aggregated contact data as contacts having occurred at “home”, “work”, “school” or “other” settings; the CoMix data was also processed to reflect this usual categorisation. We chose to focus on “work” and “other” contacts, as Google mobility does not capture changes in school contacts and residential mobility is measured in such a way that does not allow for comparison to home contacts, i.e. it measures time spent in the home, as opposed to number of visits to homes. Additionally, it is more likely that household contacts are related more closely to household size than the number of visits.

### 2.2 Data Preparation

We included participants aged 18 and over, as children’s contacts were reported by their parents, but did not restrict contact ages. As per previous CoMix analyses (15), we excluded survey rounds six and seven because of data collection issues due to an ad-hoc change to the questionnaire resulting in fewer contacts reported for those weeks. When making predictions for out-of-sample data, we include only the first year of data available to Belgium and the Netherlands due to availability of validated data. The second year of UK data was validated and available, hence this was used as another out-of-sample data source.

To average out differences in behaviour between alternating panels, we used a two-week rolling average of “work” and “other” contacts. We reweighted the sample based on age and social class (see supplementary material for visualisations of how survey participants’ characteristics changed over time, Figures S1-S3). This was to ensure that changes in the recruitment process over the course of the survey did not influence the investigation, and to improve the generalizability of the study. We also reweighted the sample so that contacts made on weekends comprised 2/7ths of the total sampling weight. This was done as slightly fewer than 2/7ths (25%) of observations were on a Saturday or Sunday and the number of contacts on these days was generally lower. Data was reweighted using weighted means, which were done over a period of two weeks - leaving the final data in the format of fortnightly.

Occasionally, some participants reported mass contacts in extremely high numbers, such as 3000 and higher. These occasional large mass-contact events introduced substantial noise into our estimates of the mean contact rate over time. To stabilise these estimates, mass contacts were capped at 50 contacts per participant, per date, for each contact type, resulting in the reduction of 6% in the total number of “work” contacts and of 7% in the total number of “other” contacts. We capped contacts by randomly sampling 50 contacts in each contact type for each participant for each date, when the contacts were listed as ‘mass’. In other words, a random selection of contacts, of size 50, was drawn from the listed contacts for that participant on a given date, respective to the contact type. Less than 1% of participants had their “work” contacts capped and less than 0.5% of participants had their “other” contacts capped. Other analyses performed on this data implemented a similar approach and hence for consistency it was applied to this investigation (15).

### 2.3 Statistical Analysis

R version 4.2.1 was used for all analyses (20); code and data are available through Github and Zenodo (see Data Availability Statement). We conducted two broad analyses: visually comparing Google mobility to contact rates, and investigating ways to use Google mobility data to scale pre-pandemic contact rates. Through the second analysis we used four methods to scale contacts, comparing methods using Google mobility alone (naïve models) and methods built using CoMix data and Google mobility data.

#### 2.3.1 Comparing Mobility Indicators to Contacts

We first visually compared the mobility indicators to the relevant contacts (see section 3.2.1): “work” contacts were compared against the “workplaces” mobility indicator and “other” contacts against a composite “other” mobility indicator which we calculated as the mean of the “retail and recreation”, “transit stations”, and “grocery and pharmacy” mobility indicators. This composite predictor was created in order to make a direct comparison between contacts from the “other” category in CoMix, and Google mobility. Here we looked at whether contact rates rose at similar time points as when mobility indicators increased. We also looked at whether restriction periods had similar impacts on both contact rates and Google mobility indicators.

#### 2.3.2 Creating Relative Contact Rates

The aim of this investigation is to determine how to use Google mobility as a proxy for social contact rates during a pandemic, specifically by comparing four different models – two “naïve” models and two regression models – relating mobility to relative contact rates. The two naïve models are commonly encountered in the literature (11–13), while the regression approach is less commonly used (21) and the specific regression models we analyse are fitted in this paper.

We have allowed for models to include an implicit level of contacts even when mobility is zero, through the inclusion of an intercept term. However, the true level of implicit contacts is not ascertainable from this estimate as the linear model is expected to “break down” in the low mobility area. Determining the number of contacts which are made during times of ‘zero visits’ to a venue type would require specific analyses, which are not available.

The two “naïve” models are based on first principles, namely that contacts are either directly proportional to mobility (“mobility” model) or to mobility squared (“mobility squared” model). The “mobility” model assumes that given a visit is made to a particular type of venue, a person makes the same number of contacts. An example of this would be going on a date: the amount of social contact you might have during a date may not strongly depend upon how many other people are going on dates. The “mobility squared” model assumes that, given a visit is made to a particular type of venue, the number of contacts depends upon the number of other people making visits to the same type of venue. An example of this might be using public transport, where your risk might depend upon how many other people are using public transport. In other words, the “mobility” model assumes that contacts are made through coordinated activities, and the “mobility squared” model assumes that contacts are made at random with other people at the same venues.

Both the “mobility” (11) and “mobility squared” (12,13) models have been used in pandemic-era modelling studies. For each of the naïve models, a mobility value of 1 corresponds to a relative contact rate of 1, and either the mobility value or the mobility value squared is used directly as the relative contact rate. The relative contact rate is then multiplied by the pre-pandemic contact matrix measured for a given setting to produce an estimate of the during-pandemic contact matrix.

The two regression models were fitted to average contact rates for the UK as measured by CoMix, for fortnightly periods, with Google mobility as the independent variable (see Figure 2). We contrasted two regression models, one with an intercept and linear term (the “linear model”) and one with an intercept, a linear term, and a quadratic term (the “quadratic model”). Like the naïve models, the regression models produce an estimate of relative contact rates from an input of mobility, which are then used as a multiplicative factor on a pre-pandemic “baseline” contact matrix, to yield a contact matrix. The pre-pandemic “baseline” was provided by estimates for the UK as measured by the 2006 POLYMOD survey (16) - see supplementary material for a more detailed description. The naïve models were each fitted separately to “work” contacts from CoMix, using Google mobility “workplace” visits as the predictor, and to “other” contacts from CoMix, using the composite Google mobility “other” visits measure as the predictor, for the UK. Confidence intervals were calculated using the confint R command (22).

The four models obtained above were used to predict relative contact rates from Google mobility data for the UK, Belgium, and the Netherlands. To assess the performance of the four models in the context of methods people may use during an emergency when there is no contact data available, we used synthetic contact matrices (23). These matrices are available for 177 countries and therefore can be used in the majority of the countries of the world. We scaled the synthetic matrices by the relative contact rates yielded by each model. We assumed that the estimated relative contact rate we have estimated for adults can be extended to children without adjustment.

From the scaled synthetic matrices, we then calculated the dominant eigenvalues for each fortnight in the study period. In order to translate these results into something understandable, we transformed the dominant eigenvalues into “partial” reproduction numbers. This was done by comparing early (07/03/2020-13/03/2020) estimates of R_t_ (number of secondary infections generated by one infected person) from the EpiForecast work (24) and the dominant eigenvalue for the synthetic matrix for ‘all’ contacts for the UK (23). This period was used as it was the earliest available estimates of R_t_ and can be considered a baseline period where scaling of POLYMOD contacts would not be required.

This gave us a multiplicative factor to use to transform the dominant eigenvalues we calculated, into partial reproduction numbers. This multiplicative factor (f) was calculated as follows:

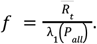

Where 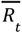 is the average of the estimates of *R*_*t*_ and λ_*t*_ (*P*_*all*_) is the dominant eigenvalue of the POLYMOD contact matrix for all contacts for the UK.

This transformation is done to put our results on a comprehensible scale, but note that in general it is not the case that partial reproduction numbers calculated in this fashion will sum to the overall reproduction number.

The “partial reproduction number” can be expressed through the following equation:

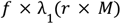

Where *r* is the ‘contact scaling factor’ derived from the regression models (detailed in supplementary section S.1) and *M* is the type-specific (e.g. work or other) synthetic contact matrix from Prem *et al*. (23) for the country of interest for which contacts are being scaled.

An alternative formulation of the “partial reproduction number”, using the linear model as an example, could be:

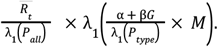

Where α + β*G* is the type-specific regression output, in which *G* is the respective mobility type, and λ_1_ (*P*_*type*_) is the dominant eigenvalue of the type-specific POLYMOD contact matrix for the UK.

We assess the accuracy of the four models in predicting partial reproduction numbers both visually (Figures 4-6) and quantitatively (Tables S6-S11), in the latter case using root mean squared error. As a ‘true’ value for the partial reproduction numbers we calculated the partial reproduction number for the CoMix contact matrices for the respective contact type and country. The same multiplicative factor was used to convert the dominant eigenvalue of the CoMix contact matrices into partial reproduction numbers, i.e. to transform type-specific contact rates into type-specific effective contacts rates.

#### 2.3.3 Work Flow Diagram

Figure 1 shows the process in which data was used throughout the investigation and how each step relates to each other. This aims to clarify how the data sources (CoMix, POLYMOD, Google Mobility) relate to one another and brings together formulae shown throughout the Methods section.

**Figure 1:**
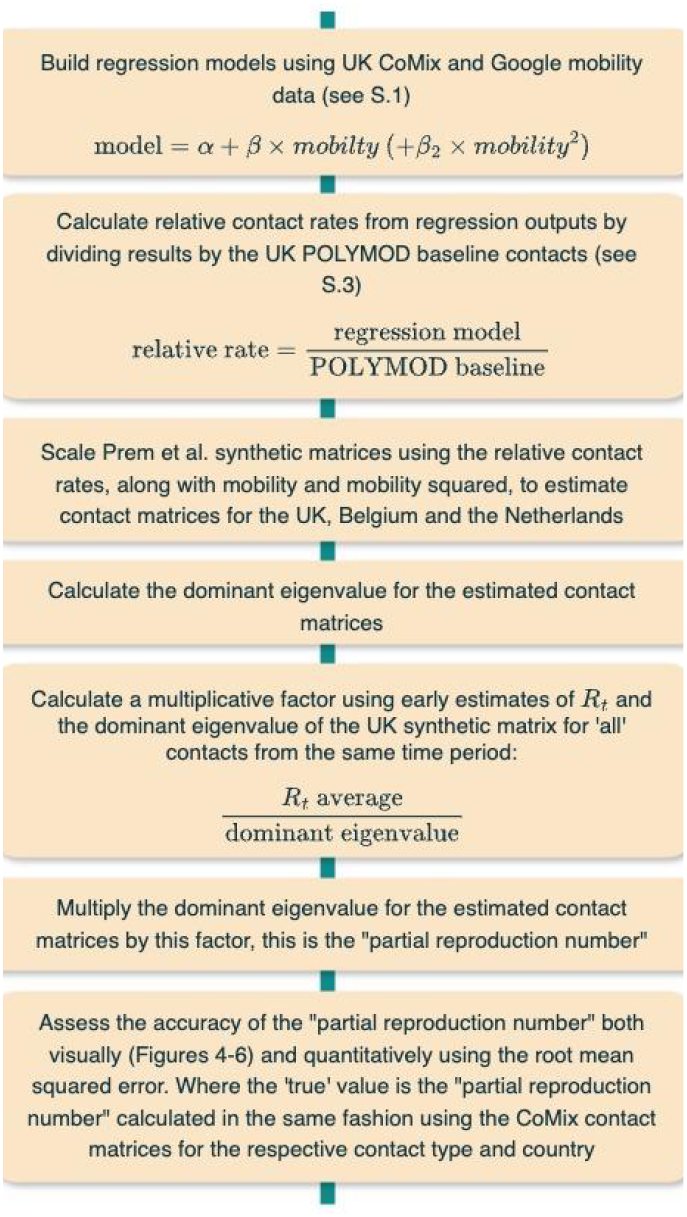
work flow diagram describing process of comparing regression based and naïve models

**Figure 2:**
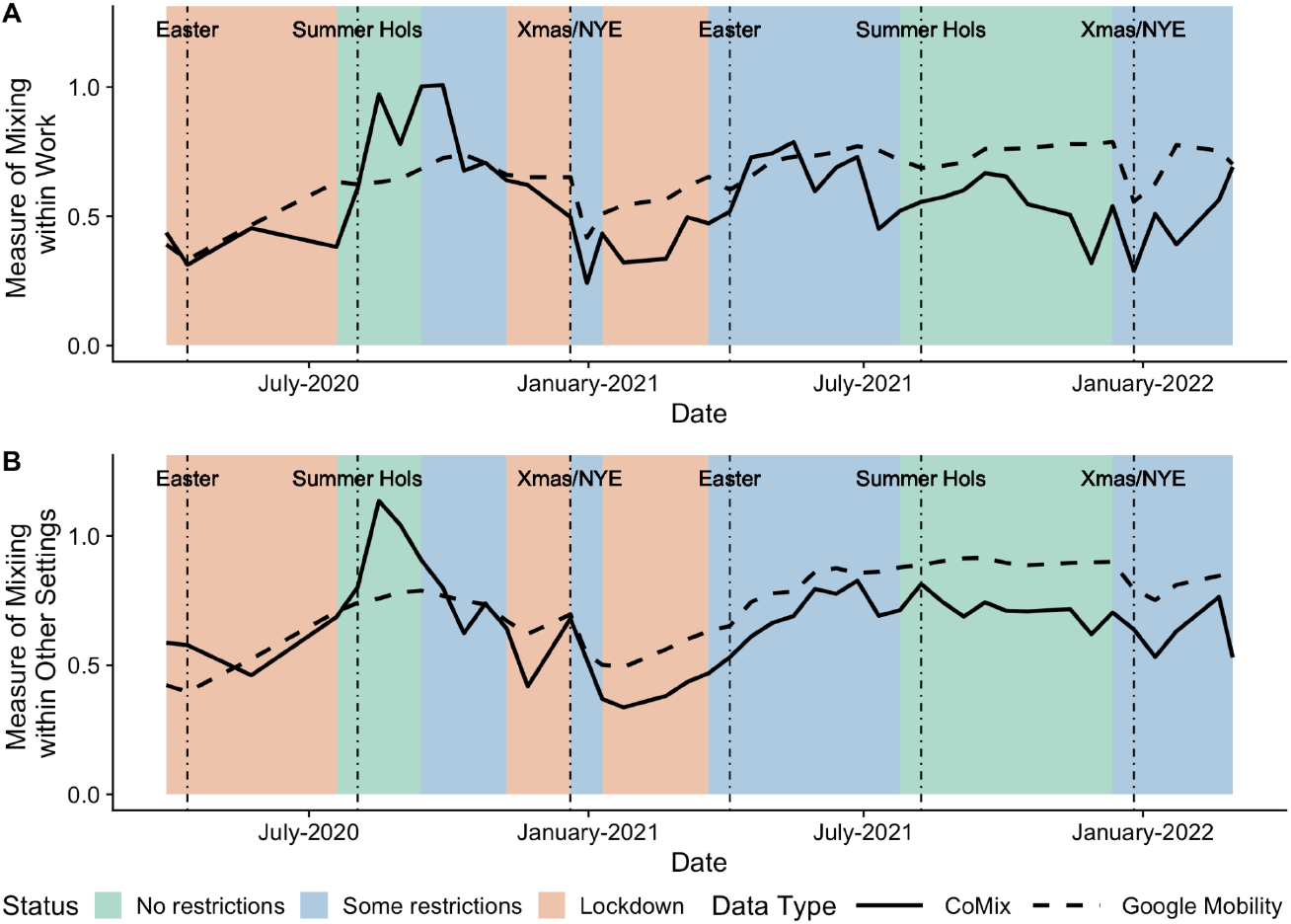
Contacts against mobility over time. Plot A shows mean “work” contacts (solid line) and “workplace” mobility (dashed line) and plot B shows mean “other” contacts (solid line) and “other” mobility (dashed line), over time respectively.

## 3. Results

### 3.1 Descriptive Analysis

For the UK there were 121,057 surveys completed in the study period, filled out by 17,497 participants. Characteristics of both the surveys completed and the individual participants are provided in Table 1. As participants were surveyed multiple times, characteristic distributions are slightly different between participants and observations.

**Table 1.** Participant characteristic summary for participants included in regression models; an observation is a complete survey response; percentages rounds to 1 decimal place so may not add to 100. Most population proportions were from the 2021 census (25) aside from social class which comes from a 2008 IPSOS social class report (26).

| *Characteristic* | *Number of Participants (%)* | *Number of Observations (%)* | *Population Proportion %* |
| --- | --- | --- | --- |
| *Gender* |  |  |  |
| Female | 8,591 (49.1) | 49,826 (50.8) | 50.75 |
| Male | 7,832 (44.8) | 45,367 (46.3) | 48.75 |
| Other | 48 (0.3) | 259 (0.3) | 0.5 |
| Missing | 1,026 (5.9) | 2,629 (2.7) | - |
| *Age-group* |  |  |  |
| 18-29 | 3,039 (17.4) | 12,552 (12.8) | 18.8 |
| 30-39 | 3,035 (17.3) | 15,401 (15.7) | 17.2 |
| 40-49 | 2,883 (16.5) | 16,279 (16.6) | 16 |
| 50-59 | 3,206 (18.3) | 19,396 (19.8) | 17.3 |
| 60-69 | 3,213 (18.4) | 20,691 (21.1) | 13.6 |
| 70+ | 2,121 (12.1) | 13,762 (14.0) | 17.2 |
| *Employment Status* |  |  |  |
| Employed | 7,508 (42.9) | 44,007 (44.9) | 45.5 |
| Unemployed | 9,989 (57.1) | 54,074 (55.1) | 54.5 |
| *Social Class* |  |  |  |
| A - upper middle class | 806 (4.6) | 4,554 (4.6) | 4 |
| B - middle class | 4,638 (26.5) | 25,820 (26.3) | 23 |
| C1 - lower middle class | 5,626 (32.2) | 32,450 (33.1) | 29 |
| C2 - skilled working class | 2,889 (16.5) | 15,561 (15.9) | 21 |
| D - working class | 2,479 (14.2) | 14,100 (14.4) | 15 |
| E - lower level of subsistence | 1,059 (6.1) | 5,596 (5.7) | 8 |
| *Area* |  |  |  |
| East Midlands | 1,350 (7.7) | 7,634 (7.8) | 7 |
| East of England | 1,664 (9.5) | 9,504 (9.7) | 9 |
| Greater London | 2,248 (12.8) | 12,889 (13.1) | 13 |
| North East | 757 (4.3) | 4,267 (4.4) | 4 |
| North West | 1,205 (6.9) | 6,571 (6.7) | 11 |
| Northern Ireland | 473 (2.7) | 2,410 (2.5) | 3 |
| Scotland | 1,592 (9.1) | 8,799 (9.0) | 8 |
| South East | 2,510 (14.3) | 13,875 (14.1) | 14 |
| South West | 1,632 (9.3) | 9,259 (9.4) | 9 |
| Wales | 922 (5.3) | 4,939 (5.0) | 5 |
| West Midlands | 1,625 (9.3) | 9,165 (9.3) | 9 |
| Yorkshire and The Humber | 1,519 (8.7) | 8,769 (8.9) | 8 |
| *Household size group* |  |  |  |
| 1 | 3,712 (21.2) | 23,211 (23.7) | 30 |
| 2 | 7,152 (40.9) | 42,874 (43.7) | 35 |
| 3-5 | 6,276 (35.9) | 30,629 (31.2) | 30 |
| 6+ | 357 (2) | 1,367 (1.4) | 5 |

Table 1 shows that overall, characteristics were broadly representative of the British population. Males were slightly underrepresented in the survey. There was also a slightly larger proportion of middle class and lower middle class participants and a slightly smaller proportion of skilled working class and working class participants compared to the population distribution. In age distribution, 60–69-year-olds were relatively overrepresented and individuals aged 70 and up were relatively underrepresented in the survey. We reweighted the sample based on participant age and social class to adjust for the most important differences.

Figure 2 compares the mean daily number of “work” contacts from CoMix to the number of visits to “workplaces” in Google Mobility, and the mean daily number of “other” contacts from CoMix to the “other” visits in Google Mobility (see methods). Trends in both data sources are broadly similar, although the relationship between mobility and contacts appears to change over time, with a notable difference between “work” contacts and “workplace” mobility in the second year of data available.

We can look closely at the periods surrounding the lockdowns in order to compare the sensitivity of both metrics, when considering large-scale changes to population behaviours.

Figure 3 shows that both metrics show similar patterns in reaction to lockdowns, with the largest changes being seen after the first lockdown. The CoMix survey began after the start of the first lockdown so we cannot compare this to pre-lockdown values, however, POLYMOD estimates of work and other contacts respectively were 1.95 and 3.48, suggesting a drastic decrease in contacts after the first lockdown. We see that for work and other, in both metrics, average contacts appear to rise after the start of the third lockdown, this is likely due to contacts recovering following the Christmas period. In addition, due to the proximity between the end of the second lockdown and the beginning of the third lockdown - and the intermediary restrictions - average contacts were already lower than other instances of pre-lockdown contacts. In addition, the sharp drop seen about a month after the implementation of the second lockdown is likely also due to changes in contacts/mobility over the Christmas period - this fact appears to be consistent across both datasets.

**Figure 3:**
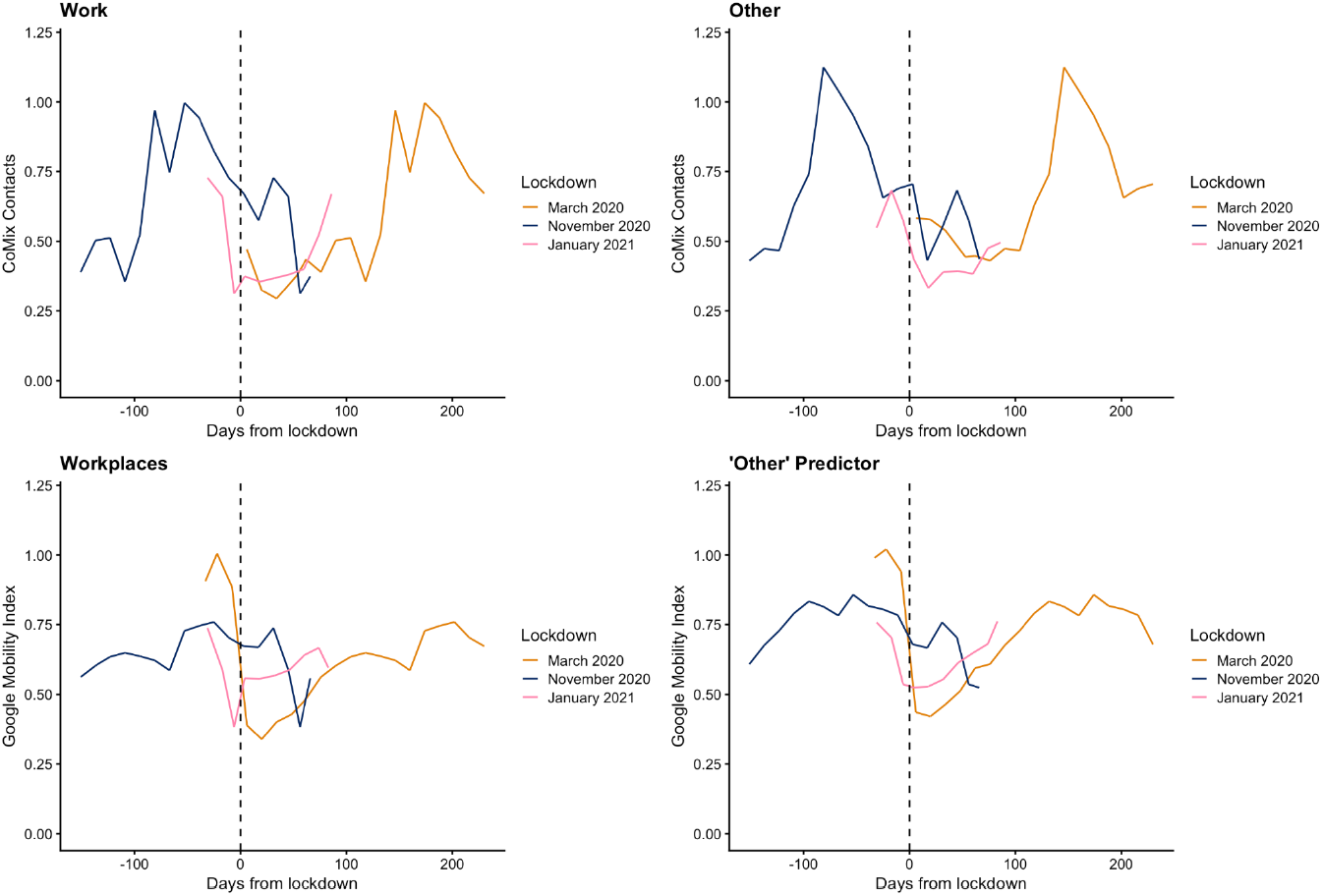
Comparison of mean contacts from CoMix survey (top row) and Google mobility data (bottom row), by number of days from respective lockdowns (1: March 2020 - orange line; 2: November 2020 - blue line; 3: January 2021 - pink line). The vertical dashed line indicates the start of the respective lockdowns.

**Figure 4:**
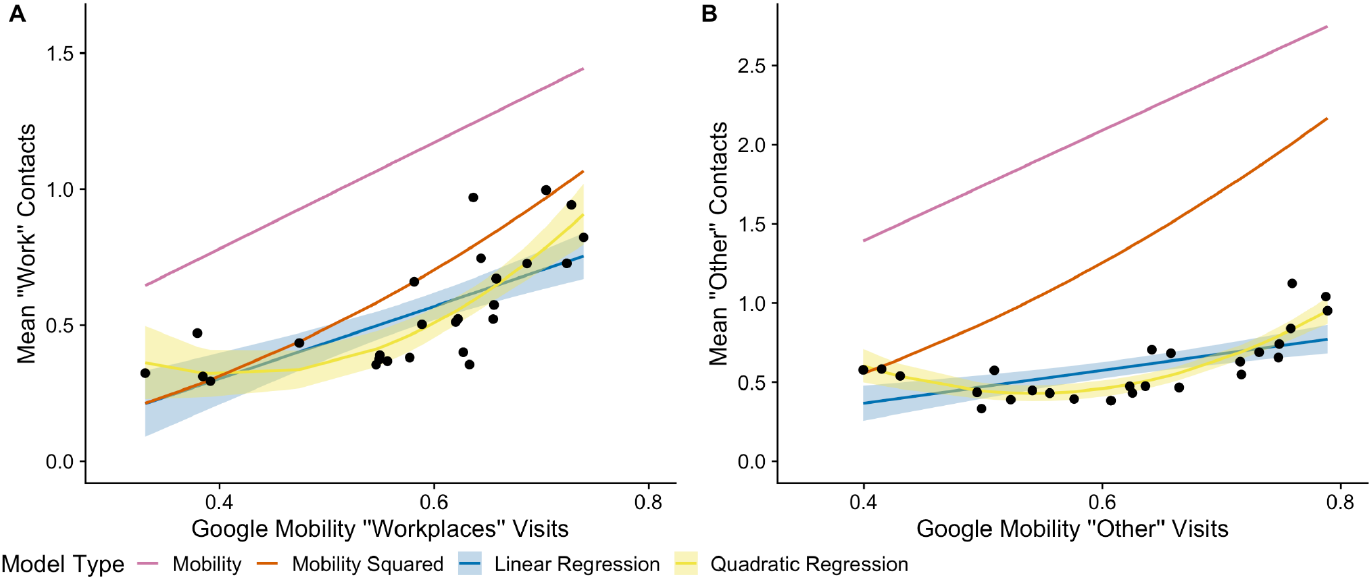
Comparison of the four models to contact data from the UK. (A) “work” contacts; (B) “other” contacts. Shaded areas show the confidence intervals for the regression model predictions and the black points show the raw data used in the regression models. The “naïve” curves are calculated by multiplying the mobility for a specific date by the POLYMOD estimate for that contact type (“work” for “workplace” mobility and “other” for “other” mobility).

In addition to comparing the mobility data to average contacts from the CoMix surveys, we also compared it to the Oxford Stringency Index (27) (supplementary figure S.4). We found that the index reflected the findings above, where the largest changes - in this case to policy - were at the onset of the first lockdown. Stringency of policy remained largely the same surrounding all other lockdown periods, which provides some explanation of why mobility and contact rates plateau.

### 3.2 Statistical Analysis

Here we compare four possible models for estimating contact rates based on mobility data to determine the most appropriate method to use when contact data is not directly available. We compare two naïve models (mobility and mobility squared) and two regression models (linear and quadratic), with the regression models fitted to data from the UK in the first year of the pandemic. We found that the quadratic regression model fitted the within-sample data better than the linear regression model (Fig. 3). Specifically, comparing the regression models by AIC, the quadratic model outperformed the linear model for both “work” contacts (linear regression: -32.4, quadratic regression: -42.6) and “other” contacts (linear regression: -29.9, quadratic regression: -57.8).

The two regression models greatly outperformed the naïve models with respect to the within-sample data. Both naïve models (mobility and mobility squared) overestimated the mean number of contacts in “work” and “other” settings (Fig. 4). However, the overestimation was less pronounced for the mobility squared model in the “work” setting. Parameter estimates for the regression models are given in the supplementary material (Tables S1-S4).

We then compared the four models’ performance on out-of-sample data: data from the UK during the second year of the pandemic, and data for Belgium and the Netherlands from the first year of the pandemic. In order to make the model fitted to UK data translatable to the other countries, we first rescaled the output of the four models according to the relative mean number of contacts in the target country (Belgium / Netherlands) compared to the UK, according to the POLYMOD study.

We then scaled synthetic matrices for the three countries (23) to the number of contacts over time in the “work” and “other” settings as calculated by the four mobility models, and we calculated the dominant eigenvalues of these scaled matrices. We then transformed the dominant eigenvalues into partial reproduction numbers using a multiplicative factor of 0.158, calculated using the formula given in section 2.3.2, to express the results on a comprehensible quantitative scale.

Focusing first on the UK setting, panels A and B in Figure 5 show that during the first year of predictions (using in-sample-data), the predictions made by the quadratic and linear models are not very distinguishable. This can be shown by comparing the root MSE for the linear models (“work”: 0.0649, “other”: 0.0365) and the quadratic models (“work”: 0.0626, “other”: 0.0264). When the second year of UK data is included (out-of-sample data), predictions diverge. Predictions from the quadratic model now over-estimate the partial reproduction number. This can be seen from the root MSE for the linear models (“work”: 0.5384, “other”: 0.2265), which are smaller than those for the quadratic models (“work”: 0.6970, “other”: 0.5569). The difference is both visually and quantitatively more clear for the “other” contacts. The results show that the best method to predict the partial reproduction number is the linear regression model, the worst method is using mobility, untransformed. This is shown by the root MSE for when only the first year of data is used (“work”: 0.2031, “other”: 0.3070) and when both first and second years are used (“work”: 1.4955, “other”: 2.3167). To see all root MSE values, see Tables S6 and S7)

**Figure 5:**
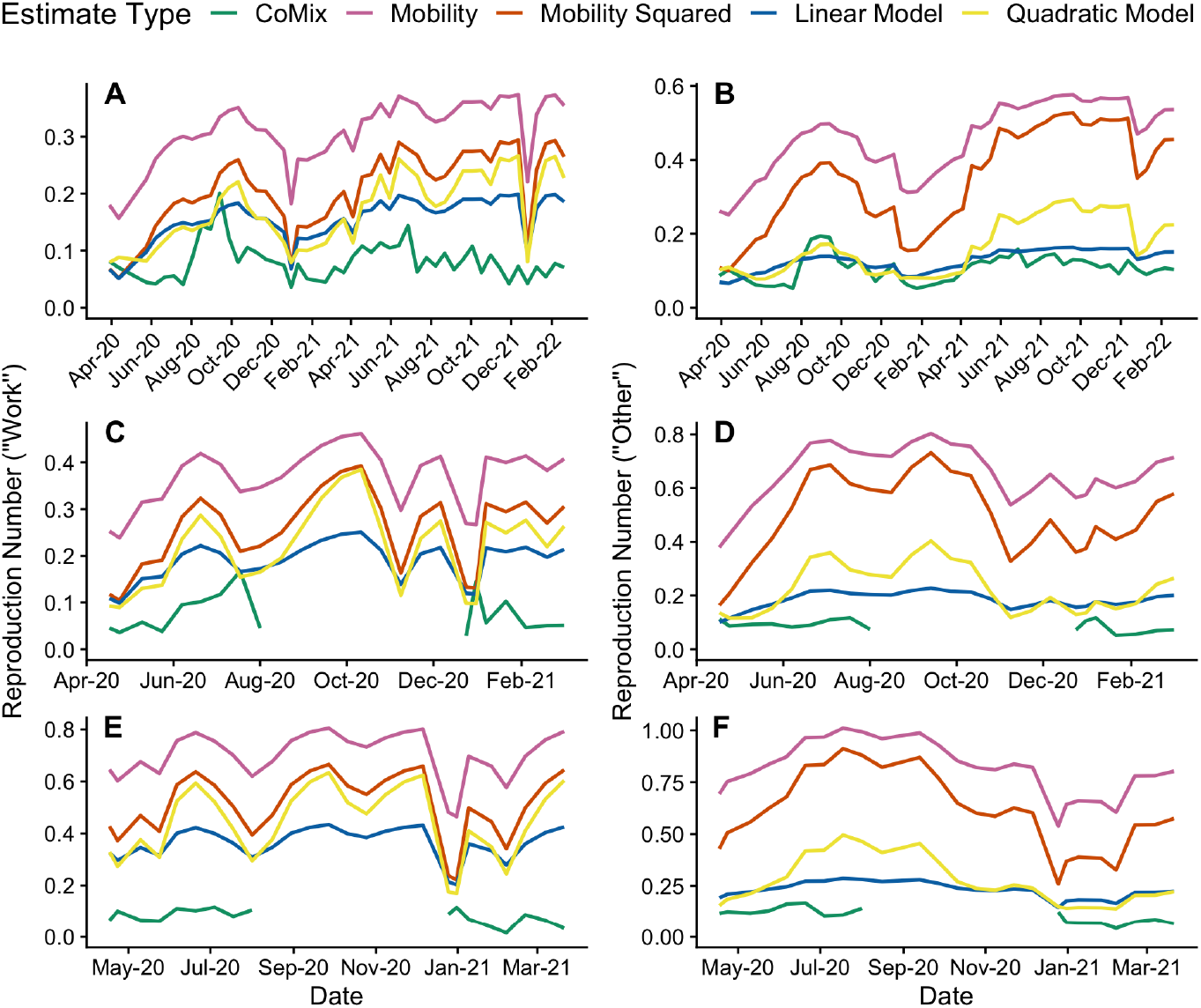
Partial reproduction numbers over time for each estimate type. Panels A and B are “work” and “other” estimates respectively, for the UK; panels C and D are “work” and “other” estimates respectively, for Belgium; panels E and F are “work” and “other” estimates respectively, for the Netherlands. For all panels the green line indicates the partial reproduction number calculated from the CoMix data; other coloured lines indicate partial reproduction numbers estimated from the four mobility models.

For Belgium, data for comparison is sparse, but where a comparison can be made, the regression model scaled estimates appear closer than the naïve model scaled estimates (panels C and D in Figure 5). When looking at the root MSE (Tables S7 and S8) the best-performing model is the linear regression model (“work”: 0.113, “other”: 0.0962) and the worst-performing is the naïve mobility model (“work”: 0.283, “other”: 0.552).

Similarly, for the Netherlands the estimates using the regression models are closer than the estimates from the naïve models (panels E and F in Figure 5). The model with the smallest root MSE was the linear regression model (“work”: 0.270, “other”: 0.121) and, as with the UK and Belgium, the model with the largest root MSE was the naïve mobility model (“work”: 0.595, “other”: 0.693).

Therefore, the linear regression model provides the most appropriate method of approximating contact rates, and subsequently “partial reproduction numbers”. When returning to first principles this suggests that the driving factor of contacts is the type of venue, as opposed to the number of other people deciding to visit this venue.

## 4. Discussion

Amidst the challenges of the COVID-19 pandemic, our research delves into the use of aggregated mobility data to understand shifting social contact rates. This investigation is the first to compare mobility indicators and contact matrices, and shows that “naive” mobility data can be improved upon as a method to measure transmission potential. We focused on predicting changes to contacts during large-scale restrictions by transforming pre-pandemic contact matrices with pandemic-era mobility data. To ascertain the most effective transformation method, we compared four approaches: two utilising regression models and two employing “naïve” models based on mobility or mobility squared. Our regression models, developed with CoMix contact survey and Google mobility data from the UK, proved more accurate in predicting contacts than the naïve models. Notably, even when applied to Belgium and the Netherlands, our regression models outperformed the naïve models.

During the COVID-19 pandemic, mobility data has been widely used to get insights into the spread of SARS-CoV-2 (28). Two main approaches have been used: some studies have tried to establish a relation between mobility and transmission (29), while others have related mobility to contact rates (30,31). While the former approach is suitable for identifying which mobility data better provides insights into epidemic spread, the latter approach has the advantage of a more natural implementation within mathematical models of infectious diseases (32,33) and is the one we focus our attention on in this work.

When comparing mobility and contact rates, we see that there was an apparent association between the respective contacts and mobility indicators (Figure 2), although this was stronger for the first year of data available. For both “work” and “other” contacts, the mean number of contacts is lower in the second year of the study relative to the corresponding mobility. This may indicate that there was a change in participant behaviour, a change in the relationship between mobility and contacts or a change in participant recruitment. This was the motivation for using the first year of data when creating the predictive models in this investigation. Indeed, the relationship between contacts and mobility is expected to be time-varying and although mobility has been found to be very predictive of social contacts during lockdown in China, this was not the case in the post-lockdown scenario (34). Also, several factors such as risk perception do affect both mobility and contacts to a different degree (35,36), therefore potentially affecting their mutual relationship.

We found that contact rates for the UK were generally better predicted, as the models used to form the relative contact rates were created using UK data. It is worth noting that CoMix data was not available for some of the study period for Belgium and the Netherlands, therefore we had less data to test the accuracy of the predictions made in these countries. This also means that the root mean squared error does not account for predictions in this period as there is no ‘true’ value to compare to. For the times where data was available for Belgium and the Netherlands, the countries were for the most part under some restrictions; this is why the CoMix estimates were often consistently low. Therefore, it is difficult to determine whether the accuracy of the approximations would be better or worse when no restrictions were in place. Nonetheless, for the UK, when restrictions were not in place, the accuracy of the approximations remained consistent with their accuracy during restriction periods.

This work aims to provide a metric which can be used to scale pre-intervention contact rates (e.g. POLYMOD, synthetic matrices, etc.) using population mobility data (e.g. Google mobility), where mobility metrics can capture changes in population behaviours when interventions are introduced/lifted. Mobility data can be combined with parameter estimates from this study (given in supplementary tables S2-S5) at any stage of an intervention (e.g. lockdown). While mobility data has limitations regarding its ability to predict contacts, it is a measure which is much more readily available when compared to contact surveys. Therefore, we offer a method which can be used to improve the accuracy with respect to using mobility data alone.

This investigation is limited by the fact that Google mobility data is difficult to define; not much information is available on how the indicators are constructed. Furthermore, Google mobility data is age-aggregated which makes it less comparable to contact data which is age-specific, and does not allow for the incorporation of potential age-specific mobility changes. In addition, CoMix surveys were not available throughout the entire study period for countries other than the UK, which limited the extent to which we were able to determine the accuracy of the results outside of the UK. As CoMix panels were surveyed repeatedly over extended periods, it is likely that a certain amount of survey fatigue began to impact responses. This is a limitation in the analysis for the UK data especially, given the number of surveys collected. In addition to this, a limitation of the study is that POLYMOD UK estimates were used to scale regression model results, in addition to informing the construction of the synthetic matrices from Prem et al., leading to potentially circular reasoning. However, the POLYMOD estimates for Belgium and the Netherlands are not used and so this limitation does not apply to estimates outside of the UK. In addition to this, we also compared the POLYMOD estimates with estimates from a survey conducted by Warwick university (37) and found both provided similar levels of baseline contacts. A further limitation of this study is that only western European countries have been used to make out-of-sample predictions, potentially limiting the applicability of our results when considering settings which may have dissimilar social behaviours or which adopted different pandemic-era restrictions. This is due to the fact that data availability of contact surveys conducted in other countries is extremely limited, and therefore there is no ‘true’ value to compare predictions to for these countries.

The study underscores the value of employing publicly-available data from pandemic contexts when estimating social contact rates without the resources for contact surveys. If given the choice between mobility and mobility squared to scale pre-pandemic contacts, the best approximation comes from mobility squared. However, the relative contact rates produced using the parameter estimates given above do provide better approximations than from using mobility data on its own. This investigation shows that the linear regression models provide the best approximations for partial reproduction number estimates out-of-sample. It is therefore our recommendation that this method is used to predict both contact matrices and reproduction numbers as opposed to using only ‘raw’ information from mobility metrics when data on social contact data is not available.

## Data Availability

CoMix survey data used for this study are available to download from Zenodo, for the UK, the Netherlands and Belgium. Google mobility data can be found on Google. The analysis code can be found on Github.

https://zenodo.org/records/13684044

https://zenodo.org/records/10549953

https://zenodo.org/records/7276465

https://www.google.com/covid19/mobility/

https://github.com/emprestige/comix_mobility

