## Supplementary Material for "Estimating social contact rates for the COVID-19 pandemic using Google mobility and pre-pandemic contact surveys"

S.1 Regression Models 2
S.1.1 “Work” contacts against “workplace” indicator 2
S.1.2 “Other” contacts against weighted predictor of indicators 2
S.2 Baseline Contact Levels 3
S.3 Relative Contact Rates 3
S.4 Supplementary Figures 4
S.4.1 Age proportions over time 4
S.4.2 Social class proportions over time 4
S.4.3 Employment status (employment age) 5
S.4.4 Oxford Stringency Index 5
S.4.5 Reproduction numbers: UK “work” contacts 6
S.4.6 Reproduction numbers: UK “other” contacts 8
S.4.7 Reproduction numbers: Belgium “work” contacts 9
S.4.8 Reproduction numbers: Belgium “other” contacts 10
S.4.9 Reproduction numbers: Netherlands “work” contacts 11
S.4.10 Reproduction numbers: Netherlands “other” contacts 12
S.5 Supplementary Tables 13
S.5.1 Comparing contacts and mobility 13
S.5.2 Regression models: “work” contacts 13
S.5.3 Regression models: “other” contacts 14
S.5.4 Reproduction numbers: UK 14
S.5.5 Reproduction numbers: BE 15
S.5.6 Reproduction numbers: NL 16
References 17

### S.1 Regression Models

#### *S.1.1 "Work" contacts against "workplace" indicator*

The linear "work" model is:

$$work = \alpha + \beta \times workplace.$$

And the quadratic "work" model is:

$$work = \alpha + \beta_1 \times workplace + \beta_2 \times workplace^2.$$

We can explicitly denote time, where $t$ is a fortnight:

The linear "work" model is:

$$work_t = \alpha + \beta \times workplace_t.$$

And the quadratic "work" model is:

$$work_t = \alpha + \beta_1 \times workplace_t + \beta_2 \times workplace_t^2.$$

#### *S.1.2 "Other" contacts against weighted predictor of indicators*

The linear "other" model is:

$$other = \alpha + \beta \times predictor.$$

And the quadratic "other" model is:

$$other = \alpha + \beta_1 \times predictor + \beta_2 \times predictor^2.$$

We can explicitly denote time, where $t$ is a fortnight:

The linear "other" model is:

$$other = \alpha + \beta \times predictor_t.$$

And the quadratic "other" model is:

$$other_t = \alpha + \beta_1 \times predictor_t + \beta_2 \times predictor_t^2.$$

### S.2 Baseline Contact Levels

As the CoMix survey was designed from the POLYMOD survey, conducted in 2005/2006 (1) this was an obvious choice for the baseline contact information. We compared this to the Warwick contact study, conducted in 2009 (2), to determine the suitability of the POLYMOD baseline. We found that both provided similar estimates of baseline contacts.

### S.3 Relative Contact Rates

For "work" contacts the relative contact rate for the linear model was calculated as follows:

$$work_{lin} = \frac{\alpha + \beta \times workplace}{baseline}.$$

And for the quadratic model:

$$work_{quad} = \frac{\alpha + \beta_1 \times workplace + \beta_2 \times workplace^2}{baseline}.$$

For "other" contacts the relative contact rate for the linear model was calculated as follows:

$$other_{lin} = \frac{\alpha + \beta \times predictor}{baseline}.$$

And for the quadratic model:

$$other_{quad} = \frac{\alpha + \beta_1 \times predictor + \beta_2 \times predictor^2}{baseline}.$$

### S.4 Supplementary Figures

#### *S.4.1 Age proportions over time*

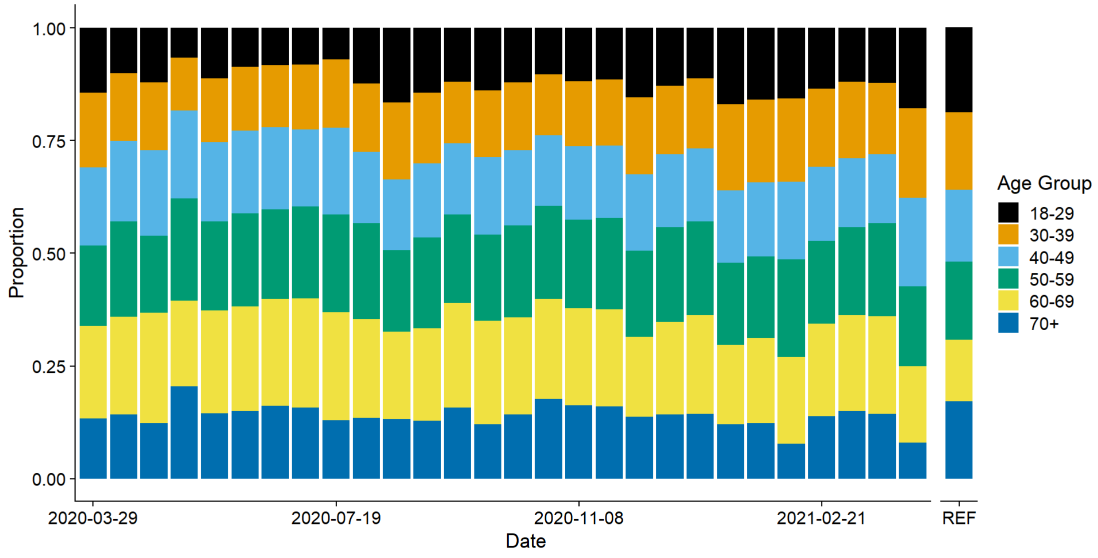


Figure S1: proportion of participants per age group by two week periods, REF shows the population proportions

#### *S.4.2 Social class proportions over time*

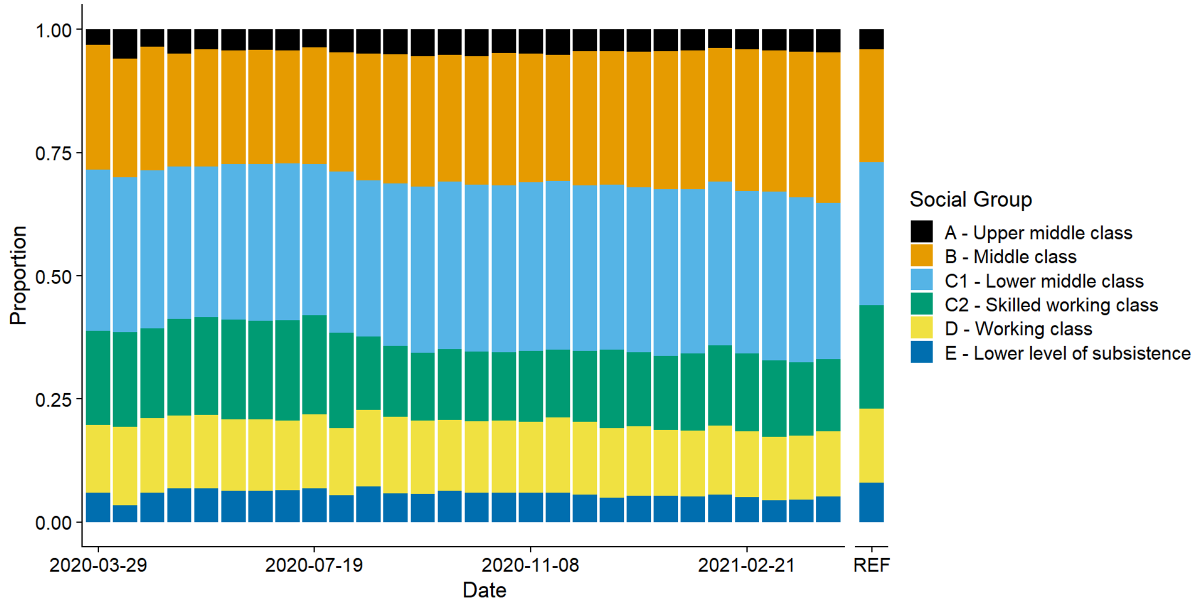


Figure S2: proportion of participants per social group by two week periods, REF shows the population proportions

#### *S.4.3 Employment status (employment age)*

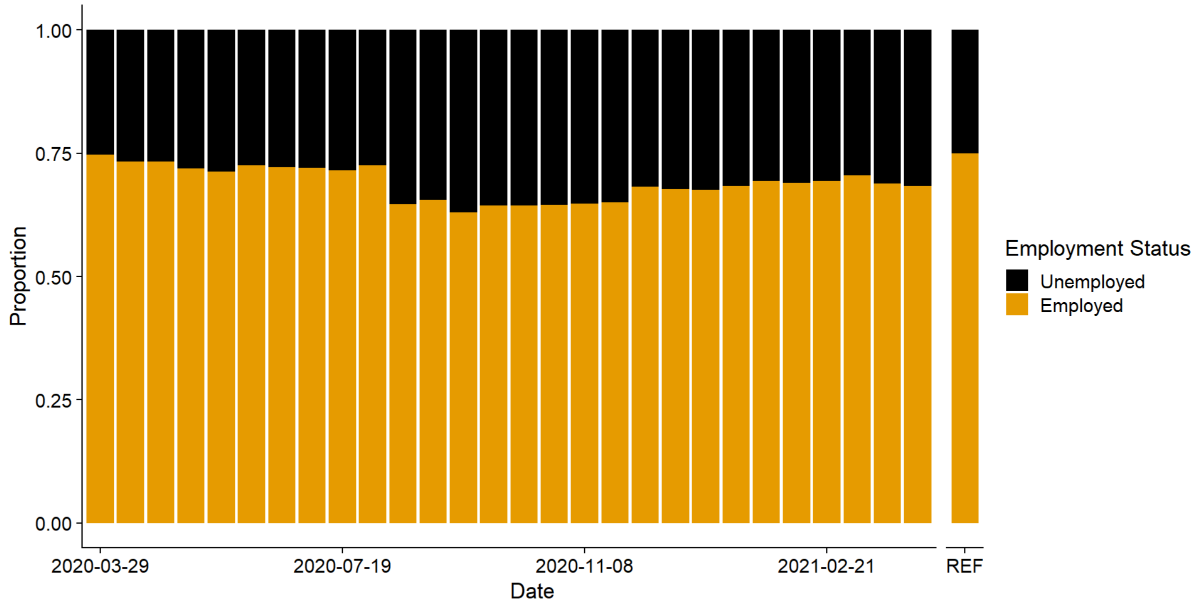


Figure S3: proportion of participants, of employment age, per employment status by two week periods, REF shows the population proportions

#### *S.4.4 Oxford Stringency Index*

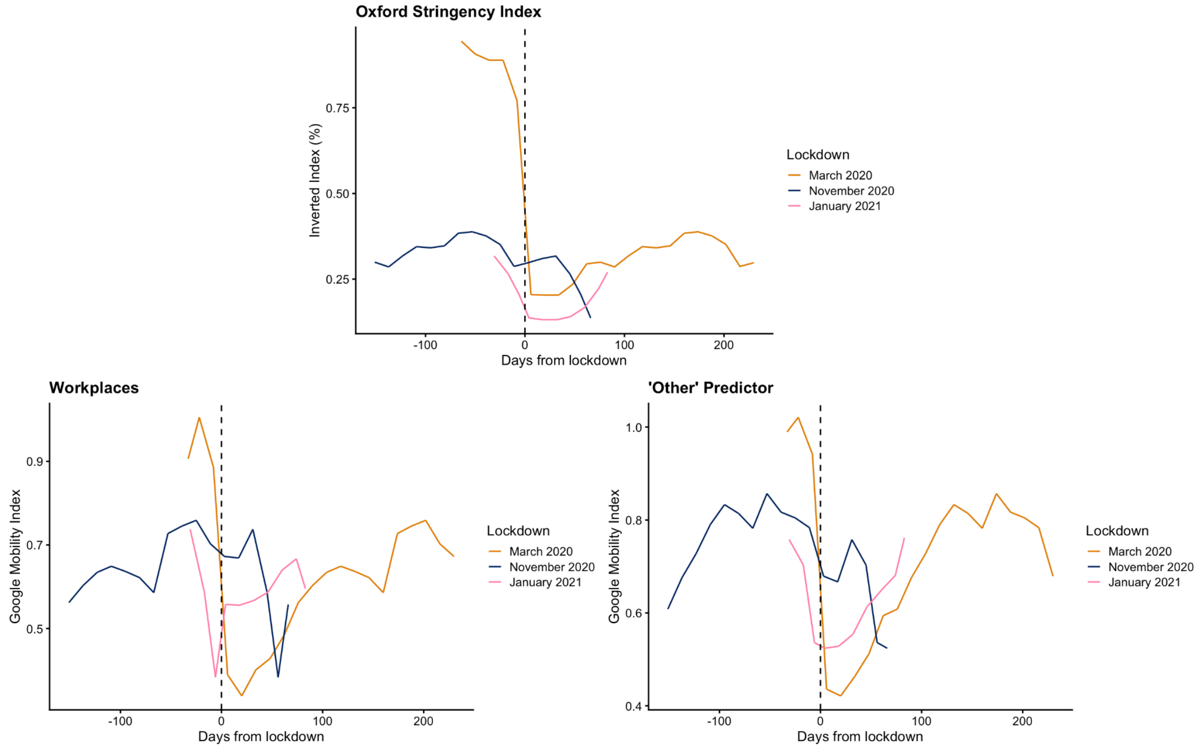


Figure S4: Comparison of the inverse of the percentage version of the Oxford Stringency Index (1 - Index/100) and Google mobility data (bottom row), by number of days from respective lockdowns (1:

March 2020 - orange line; 2: November 2020 - blue line; 3: January 2021 - pink line). The vertical dashed line indicates the start of the respective lockdowns.

*<u>S.4.5 Reproduction numbers: UK "work" contacts</u>*

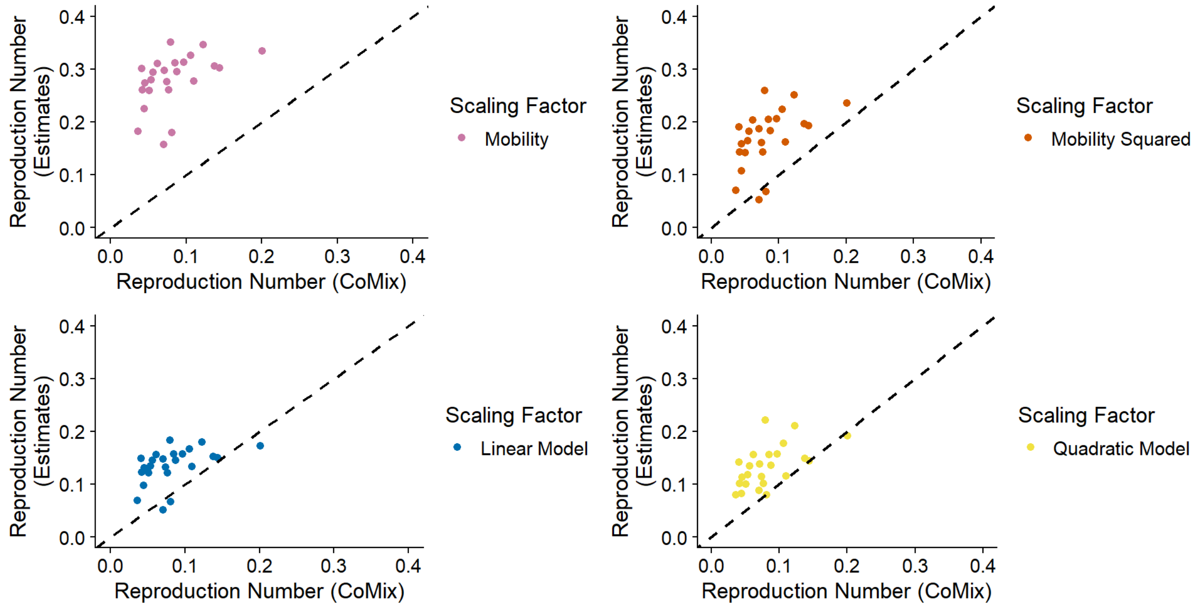


Figure S5A: reproduction number estimates against reproduction numbers from CoMix survey, top left shows estimates using mobility, top right shows estimates using mobility squared, bottom left shows estimates using the linear model relative contact rate and bottom right shows estimates using the quadratic model relative contact rate.

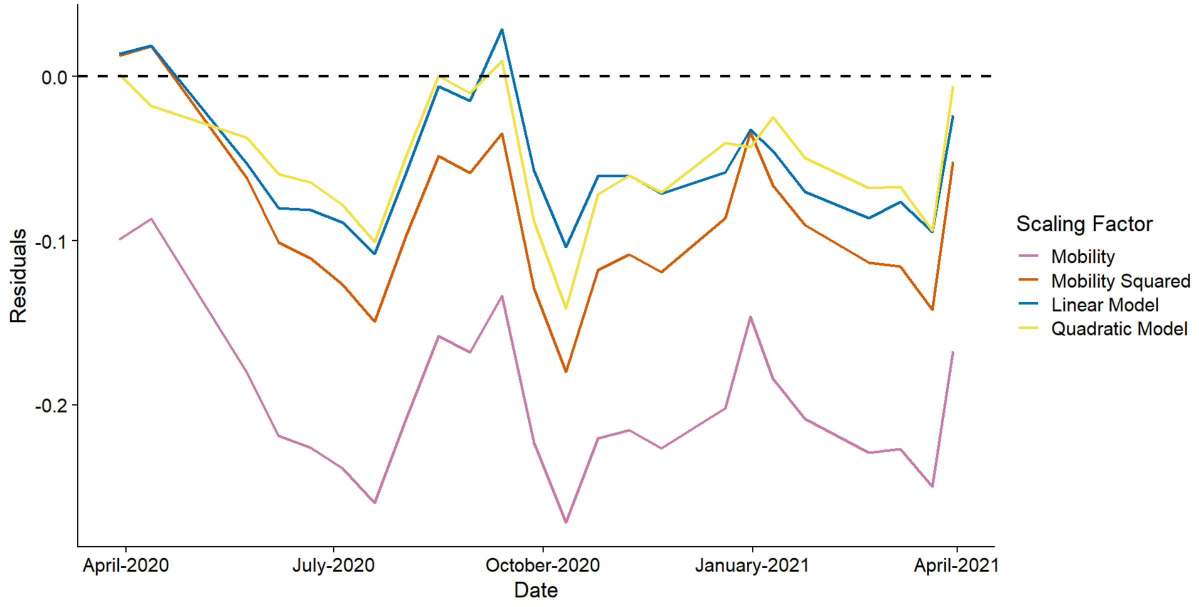


Figure S5B: residuals from the reproduction number estimates as compared to the estimates from the CoMix survey, the purple line shows estimates from the mobility scaled matrices, the red line shows

estimates from the mobility squared scaled matrices, the blue line shows estimates from the linear model scaled matrices and the orange line shows estimates from the quadratic model scaled matrices.

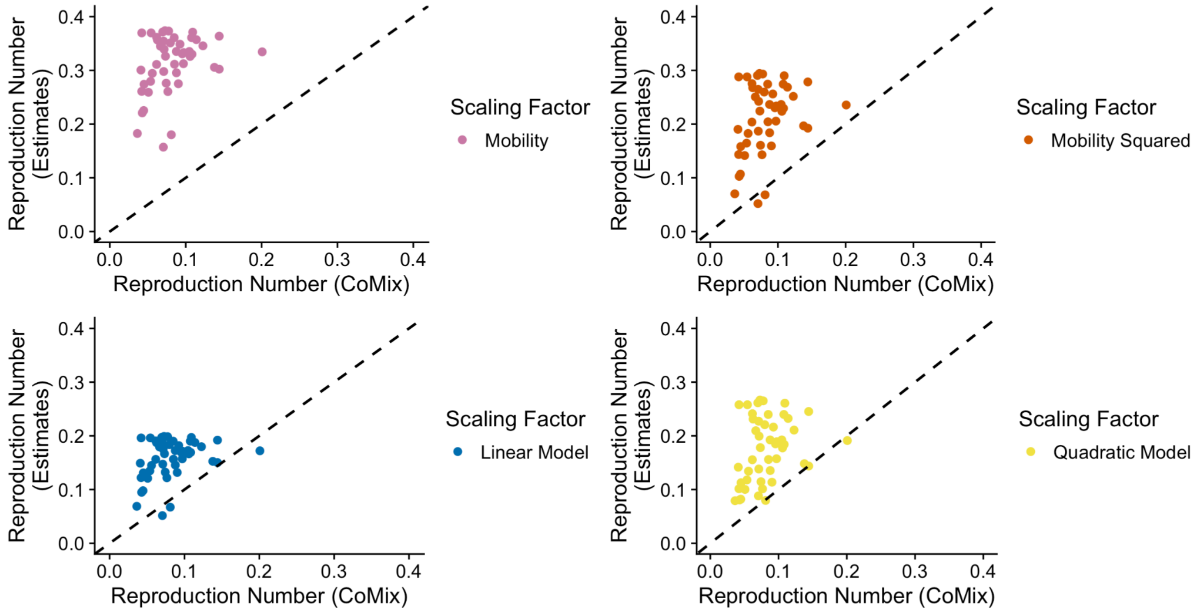


Figure S5C: reproduction number estimates against reproduction numbers from CoMix survey, for a longer time period. Top left shows estimates using mobility, top right shows estimates using mobility squared, bottom left shows estimates using the linear model relative contact rate and bottom right shows estimates using the quadratic model relative contact rate.

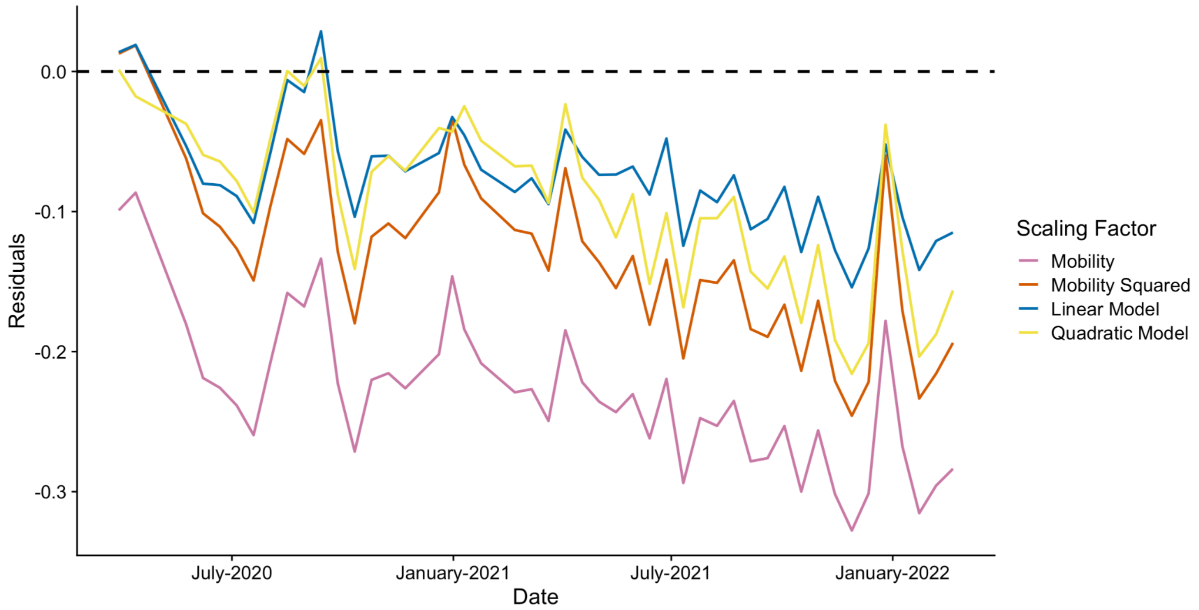


Figure S5D: residuals from the reproduction number estimates as compared to the estimates from the CoMix survey, for a longer time period. The purple line shows estimates from the mobility scaled matrices, the red line shows estimates from the mobility squared scaled matrices, the blue line shows estimates from the linear model scaled matrices and the orange line shows estimates from the quadratic model scaled matrices.

#### *S.4.6 Reproduction numbers: UK "other" contacts*

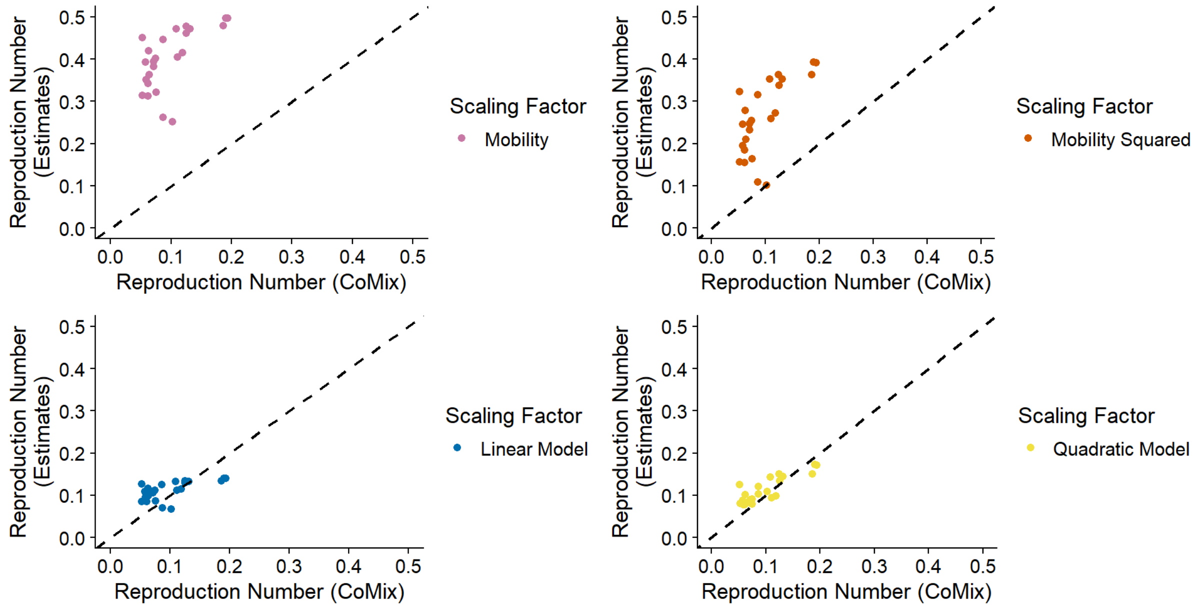


Figure S6A: reproduction number estimates against reproduction numbers from CoMix survey, top left shows estimates using mobility, for a longer time period. Top right shows estimates using mobility squared, bottom left shows estimates using the linear model relative contact rate and bottom right shows estimates using the quadratic model relative contact rate.

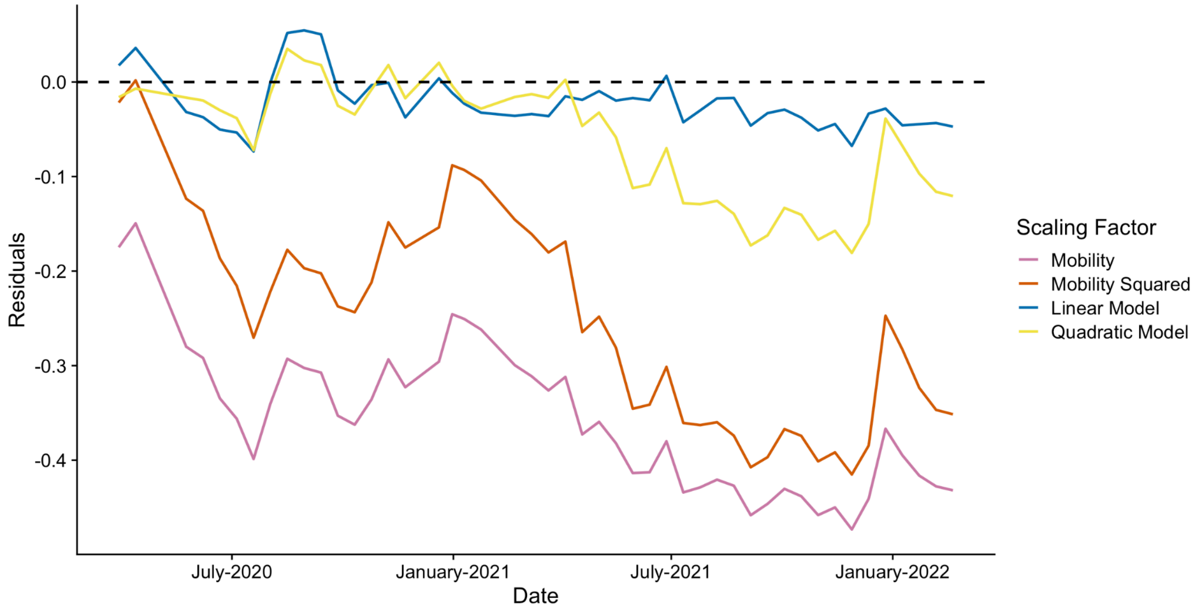


Figure S6B: residuals from the reproduction number estimates as compared to the estimates from the CoMix survey, for a longer time period. The purple line shows estimates from the mobility scaled matrices, the red line shows estimates from the mobility squared scaled matrices, the blue line shows estimates from the linear model scaled matrices and the orange line shows estimates from the quadratic model scaled matrices.

### *S.4.7 Reproduction numbers: Belgium "work" contacts*

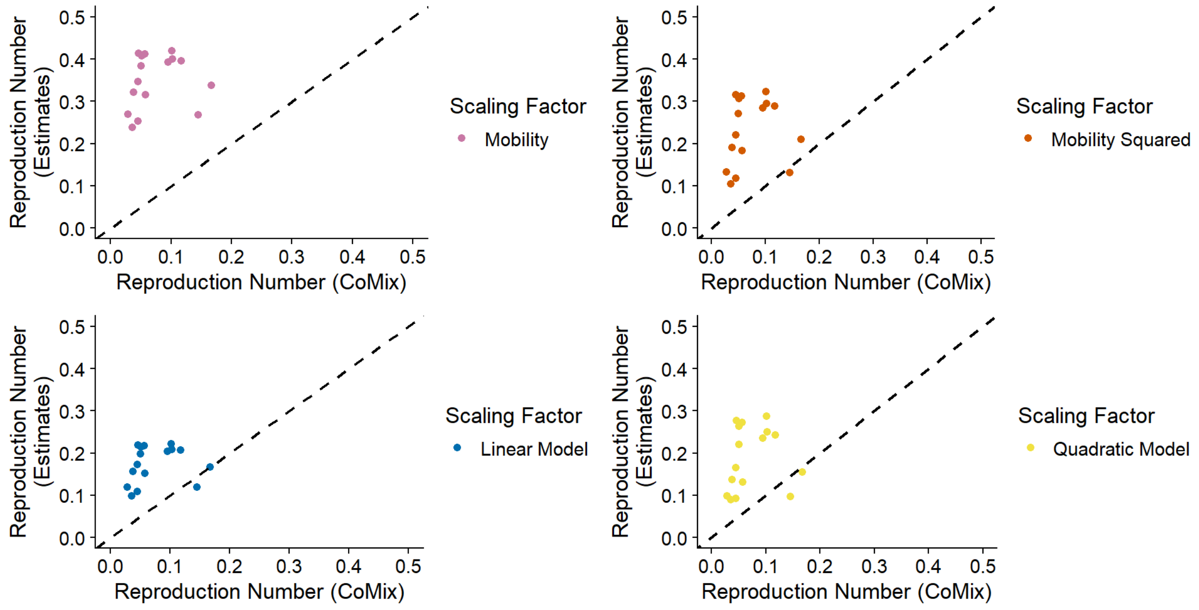


Figure S7A: reproduction number estimates against reproduction numbers from CoMix survey, top left shows estimates using mobility, top right shows estimates using mobility squared, bottom left shows estimates using the linear model relative contact rate and bottom right shows estimates using the quadratic model relative contact rate.

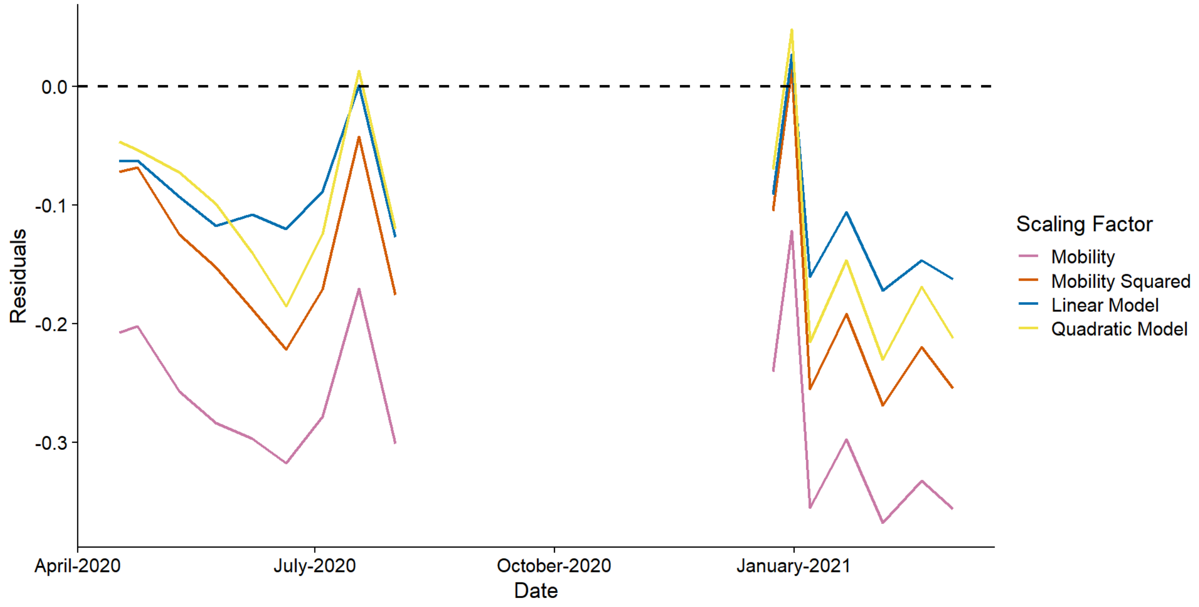


Figure S7B: residuals from the reproduction number estimates as compared to the estimates from the CoMix survey, the purple line shows estimates from the mobility scaled matrices, the red line shows

estimates from the mobility squared scaled matrices, the blue line shows estimates from the linear model scaled matrices and the orange line shows estimates from the quadratic model scaled matrices.

### *S.4.8 Reproduction numbers: Belgium "other" contacts*

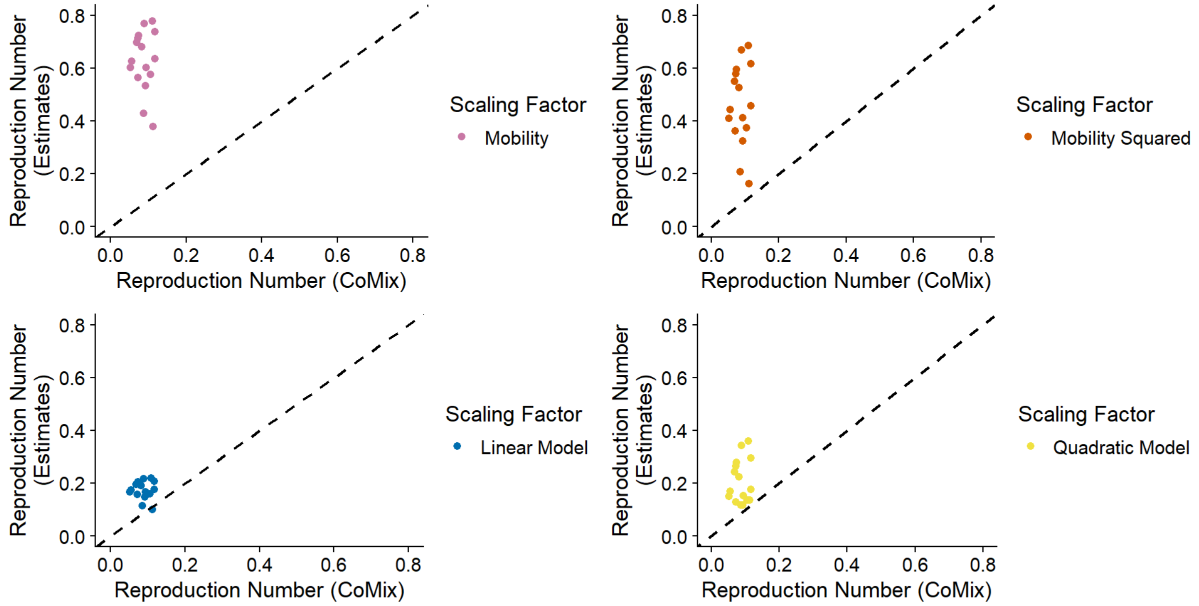


Figure S8A: reproduction number estimates against reproduction numbers from CoMix survey, top left shows estimates using mobility, top right shows estimates using mobility squared, bottom left shows estimates using the linear model relative contact rate and bottom right shows estimates using the quadratic model relative contact rate.

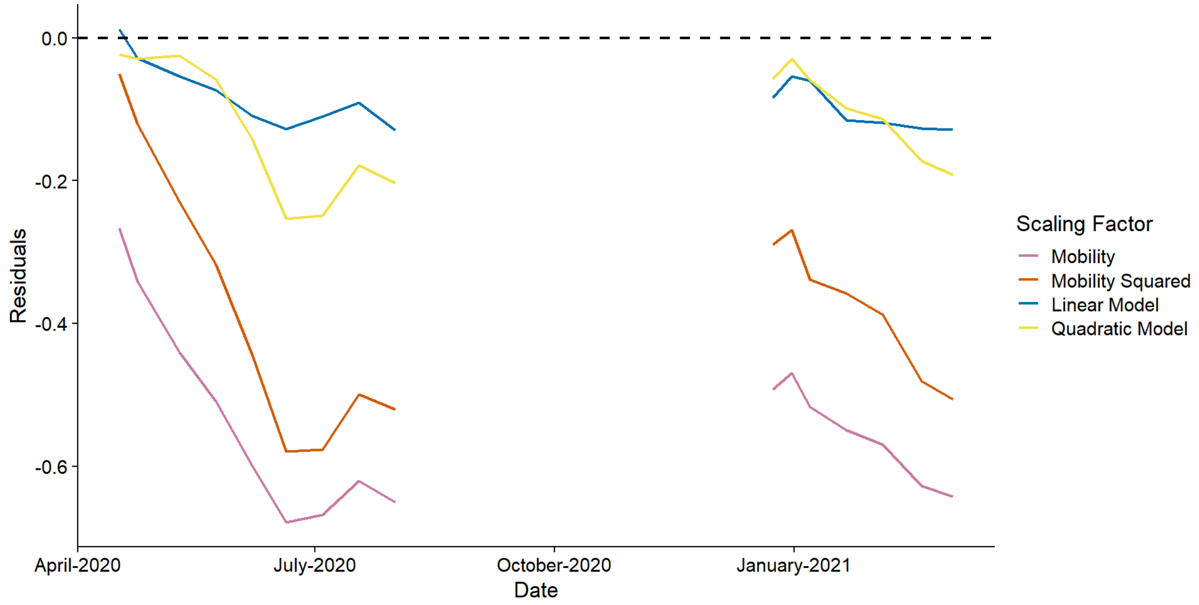


Figure S8B: residuals from the reproduction number estimates as compared to the estimates from the CoMix survey, the purple line shows estimates from the mobility scaled matrices, the red line shows

estimates from the mobility squared scaled matrices, the blue line shows estimates from the linear model scaled matrices and the orange line shows estimates from the quadratic model scaled matrices.

#### *S.4.9 Reproduction numbers: Netherlands "work" contacts*

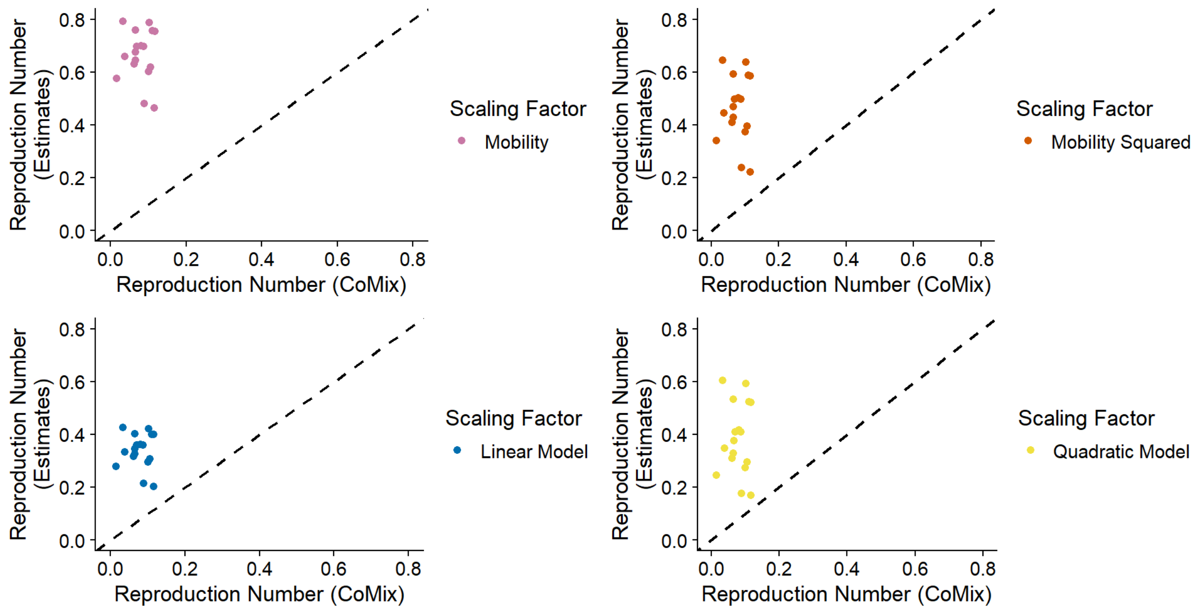


Figure S9A: reproduction number estimates against reproduction numbers from CoMix survey, top left shows estimates using mobility, top right shows estimates using mobility squared, bottom left shows estimates using the linear model relative contact rate and bottom right shows estimates using the quadratic model relative contact rate.

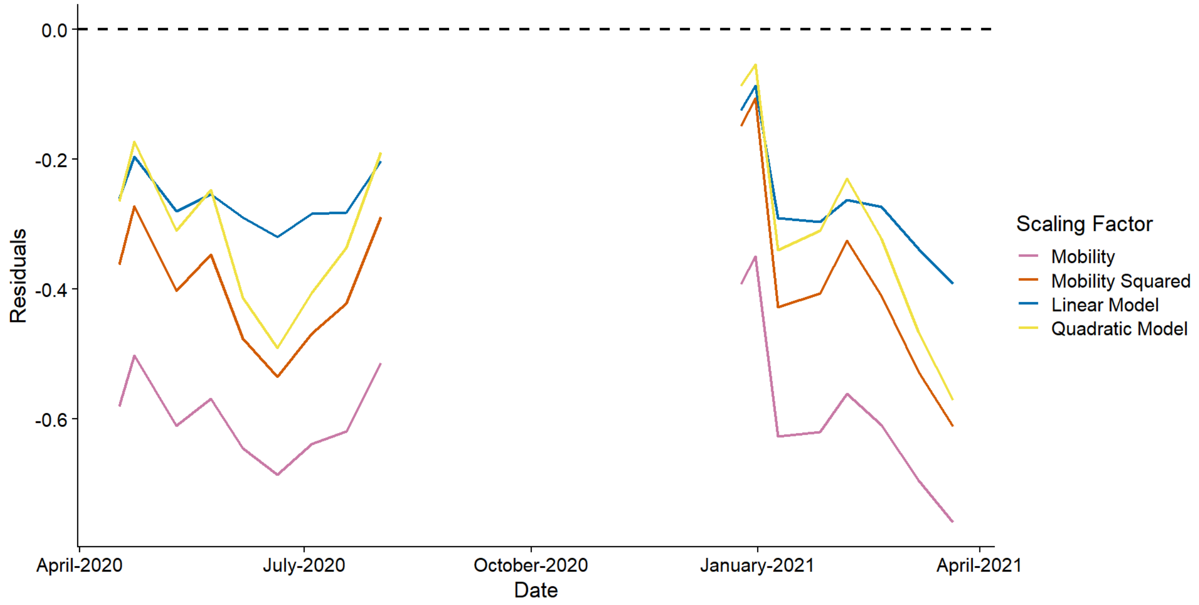


Figure S9B: residuals from the reproduction number estimates as compared to the estimates from the CoMix survey, the purple line shows estimates from the mobility scaled matrices, the red line shows

estimates from the mobility squared scaled matrices, the blue line shows estimates from the linear model scaled matrices and the orange line shows estimates from the quadratic model scaled matrices.

#### *S.4.10 Reproduction numbers: Netherlands "other" contacts*

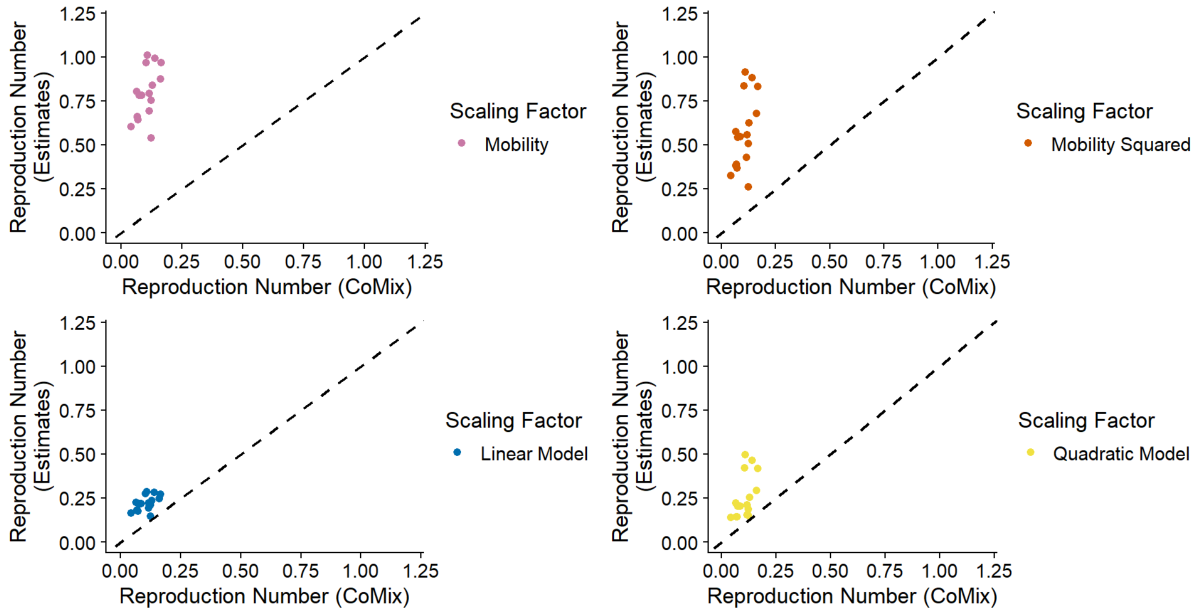


Figure S10A: reproduction number estimates against reproduction numbers from CoMix survey, top left shows estimates using mobility, top right shows estimates using mobility squared, bottom left shows estimates using the linear model relative contact rate and bottom right shows estimates using the quadratic model relative contact rate.

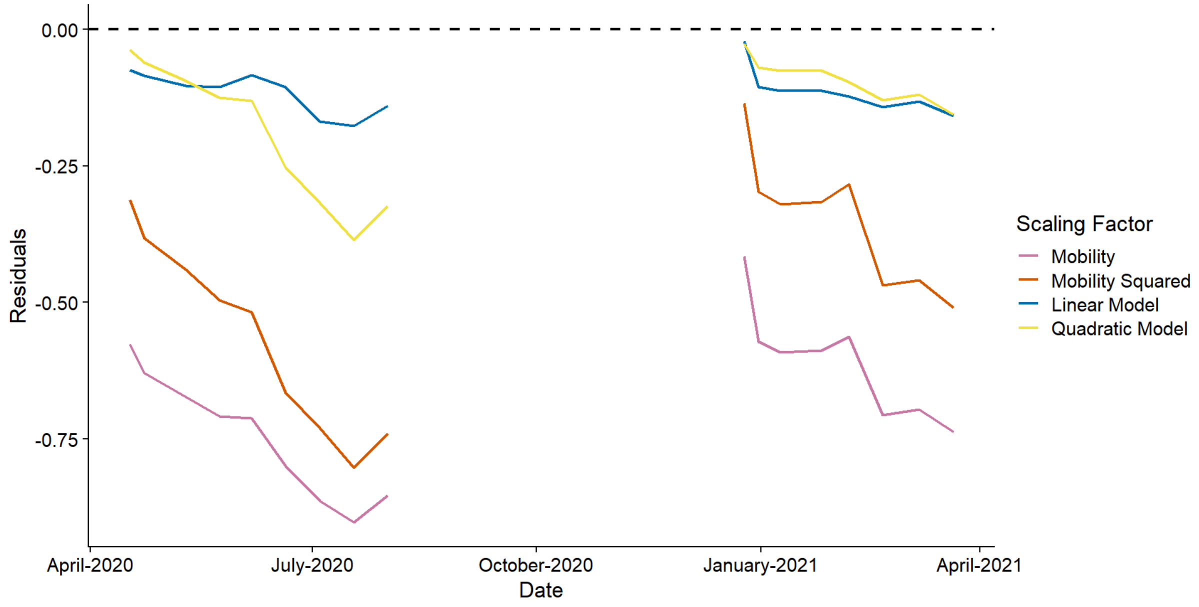


Figure S10B: residuals from the reproduction number estimates as compared to the estimates from the CoMix survey, the purple line shows estimates from the mobility scaled matrices, the red line shows

estimates from the mobility squared scaled matrices, the blue line shows estimates from the linear model scaled matrices and the orange line shows estimates from the quadratic model scaled matrices.

### S.5 Supplementary Tables

#### *S.5.1 Comparing contacts and mobility*

| *Year* | *Contact Type* | *Correlation* | *P-Value* |
| --- | --- | --- | --- |
| 1 | Work | 0.7110 | 7.461e-5 |
| 1 | Other | 0.7268 | 2.002e-5 |
| 2 | Work | 0.3222 | 0.095 |
| 2 | Other | 0.6713 | 8.154e-5 |

Table S1: correlations between contacts and mobility for the first and second years of data, Year 1 is defined as March 2020 to March 2021 and Year 2 is defined as April 2021 to April 2022. The p-values given are the result of a two-sided *t*-test.

#### *S.5.2 Regression models: "work" contacts*

| *Parameter* | *Estimate* | *95% Confidence Interval* |
| --- | --- | --- |
| Intercept | -0.2273 | (-0.4882, 0.0335) |
| "Workplace" mobility | 1.3280 | (0.8890, 1.7670) |

Table S2: parameter estimates (and confidence interval) for linear regression model for "work" contacts

| *Parameter* | *Estimate* | *95% Confidence Interval* |
| --- | --- | --- |
| Intercept | 1.3169 | (0.4275, 2.2063) |
| "Workplace" mobility | -4.7718 | (-8.1921, -1.3514) |
| "Workplace" mobility squared | 5.7062 | (2.5262, 8.8862) |

Table S3: parameter estimates (and confidence interval) for quadratic regression model for "work" contacts

*S.5.3 Regression models: "other" contacts*

| *Parameter* | *Estimate* | *95% Confidence Interval* |
|---|---|---|
| Intercept | -0.0488 | (-0.3279, 0.2304) |
| Weighted predictor | 1.0398 | (0.5971, 1.4824) |

Table S4: parameter estimates (and confidence interval) for linear regression model for "other" contacts

| *Parameter* | *Estimate* | *95% Confidence Interval* |
|---|---|---|
| Intercept | 2.9441 | (1.9893, 3.8988) |
| Weighted predictor | -9.2762 | (-12.5159, -6.0366) |
| Weighted predictor squared | 8.5566 | (5.8811, 11.2322) |

Table S5: parameter estimates (and confidence interval) for quadratic regression model for "other" contacts

*S.5.4 Reproduction numbers: UK*

| *Estimate Type* | *Root MSE* |
|---|---|
| Quadratic model | 0.0626 |
| Linear model | 0.0649 |
| Mobility squared | 0.1001 |
| Mobility | 0.2031 |
| *Including Year 2* | |
| Linear model | 0.5384 |
| Quadratic model | 0.6970 |
| Mobility squared | 0.9094 |
| Mobility | 1.4955 |

Table S6: root mean squared error for all "work" estimates for the UK, this is the square root of the mean of the squared differences between the CoMix survey reproduction number and the estimated reproduction number

| *Estimate Type* | *Root MSE* |
|---|---|
| Quadratic model | 0.0264 |
| Linear model | 0.0365 |
| Mobility squared | 0.1762 |
| Mobility | 0.3070 |
| *Including Year 2* | |
| Linear model | 0.2265 |
| Quadratic model | 0.5569 |
| Mobility squared | 1.7282 |
| Mobility | 2.3167 |

Table S7: root mean squared error for all “other” estimates for the UK, this is the square root of the mean of the squared differences between the CoMix survey reproduction number and the estimated reproduction number

#### *S.5.5 Reproduction numbers: BE*

| *Estimate Type* | *Root MSE* |
|---|---|
| Linear model | 0.1130 |
| Quadratic model | 0.1384 |
| Mobility squared | 0.1757 |
| Mobility | 0.2825 |

Table S8: root mean squared error for all “work” estimates for Belgium, this is the square root of the mean of the squared differences between the CoMix survey reproduction number and the estimated reproduction number

| *Estimate Type* | *Root MSE* |
|---|---|
| Linear model | 0.0962 |
| Quadratic model | 0.1420 |
| Mobility squared | 0.4025 |
| Mobility | 0.5520 |

Table S9: root mean squared error for all "other" estimates for Belgium, this is the square root of the mean of the squared differences between the CoMix survey reproduction number and the estimated reproduction number

#### *S.5.6 Reproduction numbers: NL*

| *Estimate Type* | *Root MSE* |
|---|---|
| Linear model | 0.2703 |
| Quadratic model | 0.3346 |
| Mobility squared | 0.4050 |
| Mobility | 0.5950 |

Table S10: root mean squared error for all "work" estimates for the Netherland, this is the square root of the mean of the squared differences between the CoMix survey reproduction number and the estimated reproduction number

| *Estimate Type* | *Root MSE* |
|---|---|
| Linear model | 0.1205 |
| Quadratic model | 0.1793 |
| Mobility squared | 0.4971 |
| Mobility | 0.6927 |

Table S11: root mean squared error for all "other" estimates for the Netherland, this is the square root of the mean of the squared differences between the CoMix survey reproduction number and the estimated reproduction number
